# Abnormal breathlessness during cardiopulmonary exercise testing - validation in people with chronic airflow limitation

**DOI:** 10.1101/2023.09.11.23295241

**Authors:** Magnus Ekström, Pei Zhi Li, Hayley Lewthwaite, Jean Bourbeau, Wan C. Tan, Dennis Jensen, the CanCOLD Collaborative Research Group

## Abstract

**Background:** Exertional breathlessness is the cardinal symptom in cardiorespiratory disease. We aimed to validate recently developed normative reference equations to evaluate breathlessness abnormality during cardiopulmonary exercise testing (CPET) in people with chronic airflow limitation.

**Methods:** Analysis of people aged ≥40 years with chronic airflow limitation undergoing CPET in the Canadian Cohort Obstructive Lung Disease (CanCOLD) study. Breathlessness intensity ratings (Borg 0-10 category ratio scale [CR10]) were evaluated in relation to power output (W), rate of oxygen uptake (V’O_2_), and minute ventilation (V’_E_) at peak exercise using normative reference equations as: 1) probability of breathlessness normality, defined as the predicted probability of the Borg CR10 rating among healthy references, with lower probability reflecting more severe breathlessness; and 2) presence of abnormal breathlessness, defined as a Borg CR10 intensity rating above the upper limit of normal (ULN). Validity of breathlessness severity (lower probability of normality) and abnormality (>ULN) was evaluated as correlations with relevant participant-reported and physiologic outcomes.

**Results:** We included 330 participants (44% women): mean±SD age 64±10 years (range 40– 89), FEV_1_/FVC 57.3±8.2%, FEV_1_ 75.6±17.9%predicted. Relative to peak W, V’O_2_ and V’_E_, abnormal breathlessness was present in 22.7%, 21.5%, and 15.2% of participants, respectively. For all equations, people with abnormal breathlessness had worse lung function, exercise capacity, self-reported symptom burden, physical activity, health-related quality of life, and physiological abnormalities during CPET.

**Conclusion:** Evaluation of breathlessness abnormality using CPET normative reference equations was valid in people with chronic airflow limitation.

## INTRODUCTION

Exertional breathlessness is a major distressing and limiting symptom in people with cardiopulmonary disease ^1^. To avoid breathlessness, many people will reduce their physical activity and enter into a vicious circle of deconditioning and further worsening of breathlessness at progressively lower levels of exertion ^2^. Given the impact of breathlessness on daily life, as well as its strong associations with adverse health outcomes including premature death ^3^, breathlessness is a major treatment target in cardiopulmonary diseases ^4^. To be able to detect abnormal breathlessness and quantify its severity and response to therapy, valid assessment of breathlessness at a standardized level exertion is key ^5,6^.

Cardiopulmonary exercise testing (CPET) is the gold standard for assessing exertional breathlessness in clinical care and research ^6-8^. Recently, normative reference equations were published for assessing abnormality of the breathlessness response to CPET, in relation to power output (W), rate of oxygen uptake (V’O_2_), and minute ventilation (V’_E_) ^9^. Each of the physiologic variables (W, V’O_2_, V’_E_) can be analyzed as absolute values or percent of the person’s predicted maximum value (%pred_max_). The normative reference equations can be used to calculate two key breathlessness measures: 1) *probability of breathlessness normality*, defined as the predicted probability of observing the person’s Borg 0-10 category ratio scale (Borg CR10) breathlessness intensity rating among healthy people (reference population) with the same covariates (age, sex, body mass, and [W *or* V’O_2_ *or* V’_E_]), where a lower probability means that the intensity rating is less likely among healthy and, thus, reflects more abnormal (or severe) exertional breathlessness; and 2) *presence of abnormal breathlessness,* defined as a Borg CR10 intensity rating above the person’s predicted upper limit of normal (ULN) ^9^.

The objective of this study was to, for the first time, validate the normative reference equations of exertional breathlessness intensity in people with chronic airflow limitation. The specific objective was to test the hypothesis that (i) probability of breathlessness normality and (ii) presence of abnormal breathlessness (>ULN) at the symptom-limited peak of incremental cycle CPET would differ by severity of airflow limitation (discriminant validity) and correlate with relevant outcomes (concurrent validity), including lung function, self-reported symptom burden and physical activity, health-related quality of life (HrQoL), exercise capacity, and exercise physiologic abnormalities mechanistically linked to exertional breathlessness ^10-14^.

## MATERIAL AND METHODS

### Study design and population

This was an analysis of the Canadian Cohort Obstructive Lung Disease (CanCOLD) study ^15^. CanCOLD is a prospective, population-based study conducted across nine communities in Canada (ClinicalTrials.gov Identifier: NCT00920348) of noninstitutionalized people aged ≥40 years originally identified through random telephone digit dialing ^15^. All participants provided written informed consent prior to completing study assessments. The research ethics board for each participating institution approved the study protocol: UBC/ PHC Research Ethics Board, P05-006 (Vancouver); Biomedical-C Research Ethics Board, BMC-06-002 (Montreal); UHN REB, 06-0421-B (Toronto); Capital Health Research Ethics Board, CDHA-RS/2007-255 (Halifax); Conjoint Health Research Ethics Board, ID21258 (Calgary); DMED-1240-09 (Kingston); 2009519-01H (Ottawa); Bio-REB09-162 (Saskatoon); CER20459 (Quebec City).

This study is reported in accordance with the STrengthening the Reporting of OBservational studies in Epidemiology (STROBE) statement ^16^.

CanCOLD participants were included in the current study if they had: 1) chronic airflow limitation defined as a post-bronchodilator forced expired volume in 1-s to forced vital capacity ratio (FEV_1_/FVC) below the predicted lower limit of normal (< LLN) ^17,18^; 2) completed a symptom-limited incremental cycle CPET at visit 1 (baseline), without premature cessation of exercise due to a supervising physician determined adverse event or protocol error; and 3) provided an intensity rating of their perceived breathlessness at peak exercise (**Figure 1**).

**Figure 1.**
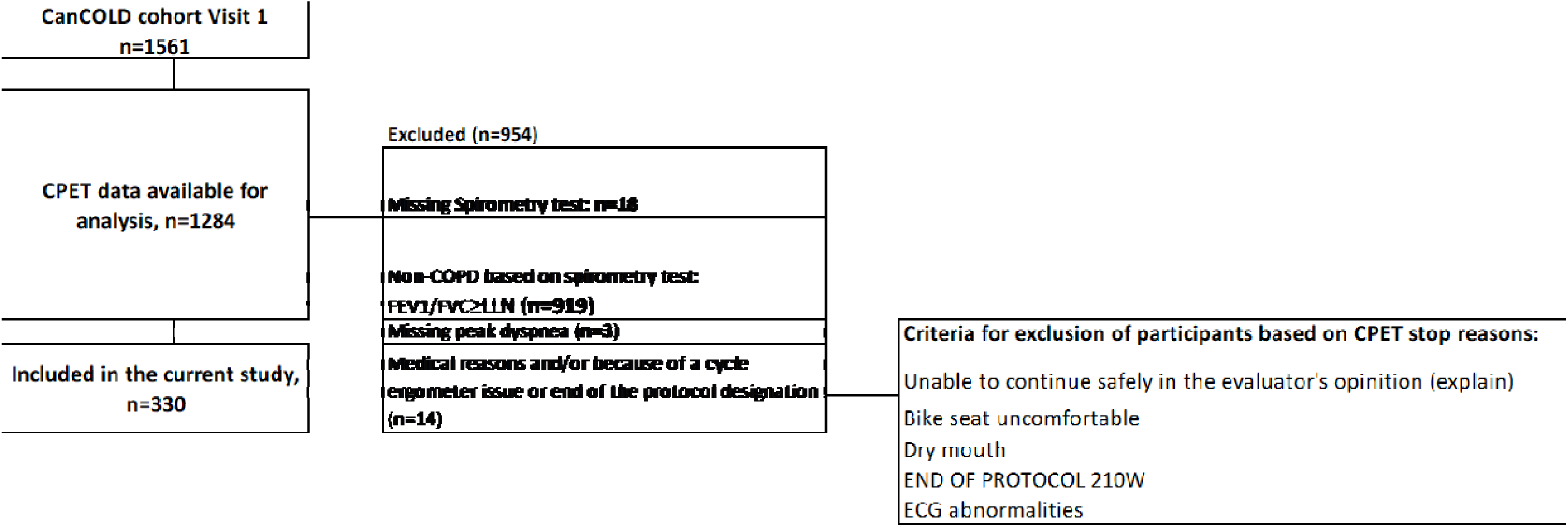
Participant flow chart.

### Assessments and procedures

All data were from CanCOLD visit 1. Participants self-reported socio-demographics and health information (e.g., smoking history and presence of physician-diagnosed health conditions) *via* structured interview with a trained researcher, whereas HrQoL was assessed using the Saint George’s Respiratory Questionnaire (SGRQ) ^19^ and COPD Assessment Test (CAT) ^20^. Impact of breathlessness on daily life activity was assessed using the Medical Research Council (MRC) 1–5 scale ^21^. Physical activity was self-reported using the Community Healthy Activities Model Program for Seniors (CHAMPS) questionnaire ^22^. Spirometry (post-bronchodilator), diffusing capacity of the lungs for carbon monoxide (D_L_CO), and plethysmographic lung volumes were assessed using automated equipment in accordance with ATS/ERS standards ^15,17,23^. Predicted lung function values were calculated using Global Lung Function Initiative (GLI) references ^24-26^. Severity of airflow limitation was categorized based on post-bronchodilator FEV_1_%predicted according to Global Initiative for Obstructive Lung Disease (GOLD) criteria: GOLD stage 1 (≥80%); GOLD stage 2 (50-79%); GOLD stage 3 (30-49%); and GOLD stage 4 (<30%) ^4^.

#### Cardiopulmonary exercise testing (CPET)

CPET was performed in accordance with recognized guidelines on an electronically braked cycle ergometer using a computerized CPET system ^27^. The CPET protocol was standardized across sites, and included a steady state pre-exercise baseline period of three to 10 minutes, followed by one minute of unloaded pedalling, and then a 10 W/min increase in power output (starting at 10 W) until symptom limitation. Participants were encouraged to maintain a pedal cadence of 50-70 rpm, and testing was stopped if pedal cadence fell below 40 rpm. Gas exchange and breathing pattern parameters were collected breath-by-breath with participants breathing through a mouthpiece and flow transducer while wearing a nose clip. Heart rate (HR) and rhythm were assessed continuously by 12-lead ECG, and peripheral oxyhemoglobin saturation (SpO_2_) was monitored by finger pulse oximetry. At rest, every two minutes during exercise, and at peak exercise, blood pressure was assessed, and participants performed maximal voluntary inspiratory capacity (IC) maneuvers ^28^, and rated the intensity (magnitude) of their perceived breathlessness and leg discomfort using the modified Borg 0-10 category ratio (CR10) scale ^29^. Prior to CPET, breathlessness was defined for each participant as “breathing discomfort” and leg discomfort as “the level of discomfort experienced during pedalling”; and participants were familiarized with Borg’s CR10 scale such that “0” represented “no breathing (leg) discomfort” and “10” represented “the most severe breathing (leg) discomfort that you have ever experienced or can imagine experiencing”.

#### Analysis of breathlessness responses

Abnormality of breathlessness was evaluated using published CanCOLD normative reference equations in relation to peak W, V’O_2_, and V’_E_, respectively ^9^. Peak W was taken as the highest power output a participant was able to sustain for ≥30-s, whereas peak V’O_2_ and V’_E_ were taken as the average of the last 30-s of loaded pedalling.

Using each of the three normative reference equations, the *probability of breathlessness normality* was defined as the predicted probability of each participant’s Borg CR10 breathlessness intensity rating at peak exercise ^9^. This probability reflects how likely each participant’s breathlessness intensity rating would be at any given peak W, V’O_2_, and V’_E_ relative to a healthy reference with similar age, sex and body mass, where a lower probability reflects a more abnormal (or severe) exertional breathlessness response ^9^. For example, a predicted probability of 0.001 would indicate that less than 1/1,000 healthy people (with similar age, sex, and body mass) would report this or a higher breathlessness intensity rating. The *presence of abnormal exertional breathlessness* was categorized as a Borg CR10 breathlessness intensity rating at peak exercise > ULN, which corresponds to a probability < 0.05 ^9^.

#### Analysis of physiological responses to CPET

Physiological variables evaluated in the current study included:

- Exercise capacity as peak W and V’O_2_;
- Change in IC from pre-exercise baseline to peak exercise expressed in litres (Δ IC) and indexed to peak V’_E_ (Δ IC/V’_E_);
- Proportion of people with clinically significant dynamic hyperinflation, defined as decrease in IC > 0.15L from pre-exercise baseline to peak exercise ^30^;
- Nadir of the ventilatory equivalent for carbon dioxide (V’_E_/V’CO_2_), identified as the lowest 30-s average data point observed during CPET;
- Proportion of people with exercise ventilatory inefficiency, defined as nadir V’_E_/V’CO_2_ >ULN, and >34 ^31^; and
- Critical inspiratory constraints as the (i) tidal volume-to-inspiratory capacity ratio (V_T_%IC) and (ii) end-inspiratory lung volume-to-total lung capacity ratio (EILV%TLC), with both ratios indexed to peak V’_E_ (V_T_%IC/V’_E_ and EILV%TLC/V’_E_).

Unless indicated otherwise, all physiological variables and Borg CR10 breathlessness intensity ratings used in the analyses were values at peak exercise, to have a common point of assessment for comparisons. Predicted values for the physiological parameters at the symptom-limited peak of CPET were calculated using CanCOLD normative reference equations ^31^.

### Statistical analyses

Participant characteristics and outcome variables (self-reported and physiological variables) were summarized using mean with standard deviation (SD) and median with range or interquartile range (IQR) for continuous variables, as appropriate. Categorical variables were expressed as frequencies and percentages. No data were imputed.

Discriminant validity of the predicted probability of breathlessness normality was analyzed by plotting the relationship between the predicted probability (categorized as: 0.74-0.50; 0.49-0.25; 0.24-0.05; and < 0.05) and each outcome variable with 95% confidence intervals (CIs).

Discriminant validity of the presence of abnormal exertional breathlessness (>ULN) *vs.* normal exertional breathlessness (≤ULN) was analyzed as the between-group difference in each outcome, using Student’s t-tests and Wilcoxon-Mann-Whitney test for continuous and Chi-square and Fishers exact test for categorical variables. To assess concurrent validity, correlations between abnormal breathlessness and each outcome was analyzed as point biserial correlations ^32^. All analyses were performed using each normative reference equation (one for each of W, V’O_2_, and V’_E_).

Statistical significance was defined as two-sided p-value < 0.05. Statistical analyses were conducted using the SAS version 9.4 software (TS1M5) (SAS Institute Inc., Cary, NC, USA, 2016).

## RESULTS

A total of 330 participants (44% women) with chronic airflow limitation were included (**Figure 1**). Participants had a mean age of 64.4 (SD 10.2) years and BMI of 27.2 kg/m^2^ (SD 4.9). The majority of participants: were relatively asymptomatic (10% and 39.4% had MRC breathlessness rating ≥3 and CAT total score ≥10, respectively); had only mild to moderate airflow chronic limitation (41% had GOLD stage 1, and 51% GOLD stage 2); did not have a prior physician-diagnosis of COPD (38.5%); and were not taking any respiratory medication(s) at the time of CanCOLD visit 1 (49.7%) (**Table 1**).

**Table 1.** Participant characteristics.

| Characteristic | All (n=330) |
| --- | --- |
| Age, years, mean (SD) [min, max] | 64.4 (10.2), 64.0 (40.0, 89.0) |
| Women, n (%) | 145 (43.9) |
| Body mass, kg | 77.9 (16.3) |
| Body mass index, kg/m<sup>2</sup> | 27.2 (4.9) |
| Cigarette smoking status, n (%) |  |
| Never smoker | 93 (28.2) |
| Former smoker | 165 (50.0) |
| Current smoker | 72 (21.8) |
| Cigarette smoker pack-years | 25.8 (24.9) |
| Hypertension, n (%) | 104 (31.5) |
| Physician-diagnosed COPD, n (%) | 127 (38.5) |
| Physician-diagnosed asthma, n (%) | 139 (42.1) |
| Any respiratory medication(s), n (%) | 164 (49.7) |
| MRC breathlessness rating | 1.7 (0.7) |
| 1, n (%) | 150 (45.5) |
| 2, n (%) | 136 (41.2) |
| ≥3, n (%) | 33 (10.0) |
| HADS anxiety score | 4.0 (3.1) |
| HADS depression score | 2.9 (2.7) |
| CAT | 8.9 (6.8) |
| CAT≥10, n (%) | 130 (39.4) |
| SGRQ-Total score | 18.5 (16.1) |
| CHAMPS moderate and greater intensity, Caloric expenditure per week (KC) | 2330.9 (2452.3) |
| CHAMPS all activities, Caloric expenditure per week (KC) | 4093.8 (2983.7) |
| **Lung function** |  |
| FEV<sub>1</sub>, %pred | 75.6 (17.9) |
| GOLD stage, n (%) |  |
| GOLD 1 (FEV<sub>1</sub>/FVC <0.7 and FEV<sub>1</sub> ≥ 80% predicted) | 135 (40.9) |
| GOLD 2 (FEV<sub>1</sub>/FVC <0.7 and FEV<sub>1</sub> ≥ 50% and < 80% predicted) | 167 (50.6) |
| GOLD 3 (FEV<sub>1</sub>/FVC <0.7 and FEV<sub>1</sub> ≥ 30% and < 50% predicted) | 27 (8.2) |
| GOLD 4 (FEV<sub>1</sub>/FVC <0.7 and FEV<sub>1</sub> < 30% predicted) | 1 (0.3) |
| FVC, %pred | 102.1 (19.4) |
| FEV<sub>1</sub>/FVC, % | 57.3 (8.2) |
| TLC, %pred | 110.2 (15.3) |
| IC, %pred | 95.1 (22.7) |
| D<sub>L</sub>CO, %pred | 87.9 (23.5) |
| **Symptom-limited peak CPET parameters** |  |
| Power output (W), %pred | 81.5 (25.2) |
| V'O<sub>2</sub>, %pred | 82.7 (23.1) |
| V'E, %pred | 83.5 (24.7) |
| Nadir V'E/V'CO<sub>2</sub> | 32.0 (6.7) |
| V'E/V'CO<sub>2</sub> nadir > 34, n (%) | 100 (30.4) |
| $V'_E/V'CO_2$ nadir > ULN, n (%) | 76 (23.1) |
| $V_T\%IC/V'_E$ | 1.41 (0.53) |
| $EILV\%TLC/V'_E$ | 1.81 (0.73) |
| $\Delta IC$ (L) | -0.22 (0.39) |
| Decrease in IC > 0.15L from baseline to peak exercise, n (%) | 173 (56.0) |
| $\Delta IC/V'_E$ | -0.004 (0.009) |
| Breathlessness (Borg CR10), median (Q1, Q3) | 5.0 (4.0,7.0) |
| Leg discomfort (Borg CR10), median (Q1, Q3) | 7.0 (4.0,8.0) |

Data presented as mean (standard deviation) unless otherwise specified.
*Abbreviations:* CAT = COPD Assessment Test; CHAMPS = Community Healthy Activities Model Program for Seniors questionnaire; COPD = chronic obstructive pulmonary disease; CR = category-ratio; DBP = diastolic blood pressure; $D_LCO$ = diffusion lung capacity for carbon monoxide; EILV = end-inspiratory lung volume; $FEV_1$ = forced expired volume in one second; FVC = forced vital capacity; GOLD = Global Initiative for Obstructive Lung Disease; HADS = Hospital Anxiety and Depression Scale; HR = heart rate; IC = inspiratory capacity; MRC = Medical Research Council; RER = respiratory exchange ratio; RV = residual volume; SBP = systolic blood pressure; SD = standard deviation; SGRQ = Saint George's Respiratory Questionnaire; $SpO_2$ = peripheral saturation of oxygen; TLC = total lung capacity; Q = quartile; $V'CO_2$ = rate of exhaled carbon dioxide; $V'_E$ = minute ventilation; $V'O_2$ = rate of oxygen uptake; $V_T$ = tidal volume; W = watt; $\Delta$ = change from baseline to peak exercise.

### Probability of breathlessness normality

The distribution of predicted probabilities for the peak breathlessness intensity ratings relative to W, V’O_2_ and V’_E_ are shown in **Figure S1**. A lower probability of breathlessness normality (reflecting more abnormal [or severe] exertional breathlessness) was related to having worse outcomes, including: lower lung function; greater self-reported symptom burden; lower self-reported physical activity; lower HrQoL; lower exercise capacity; and more severe physiological abnormalities at peak exercise (**Figure 2**).

**Figure 2.**
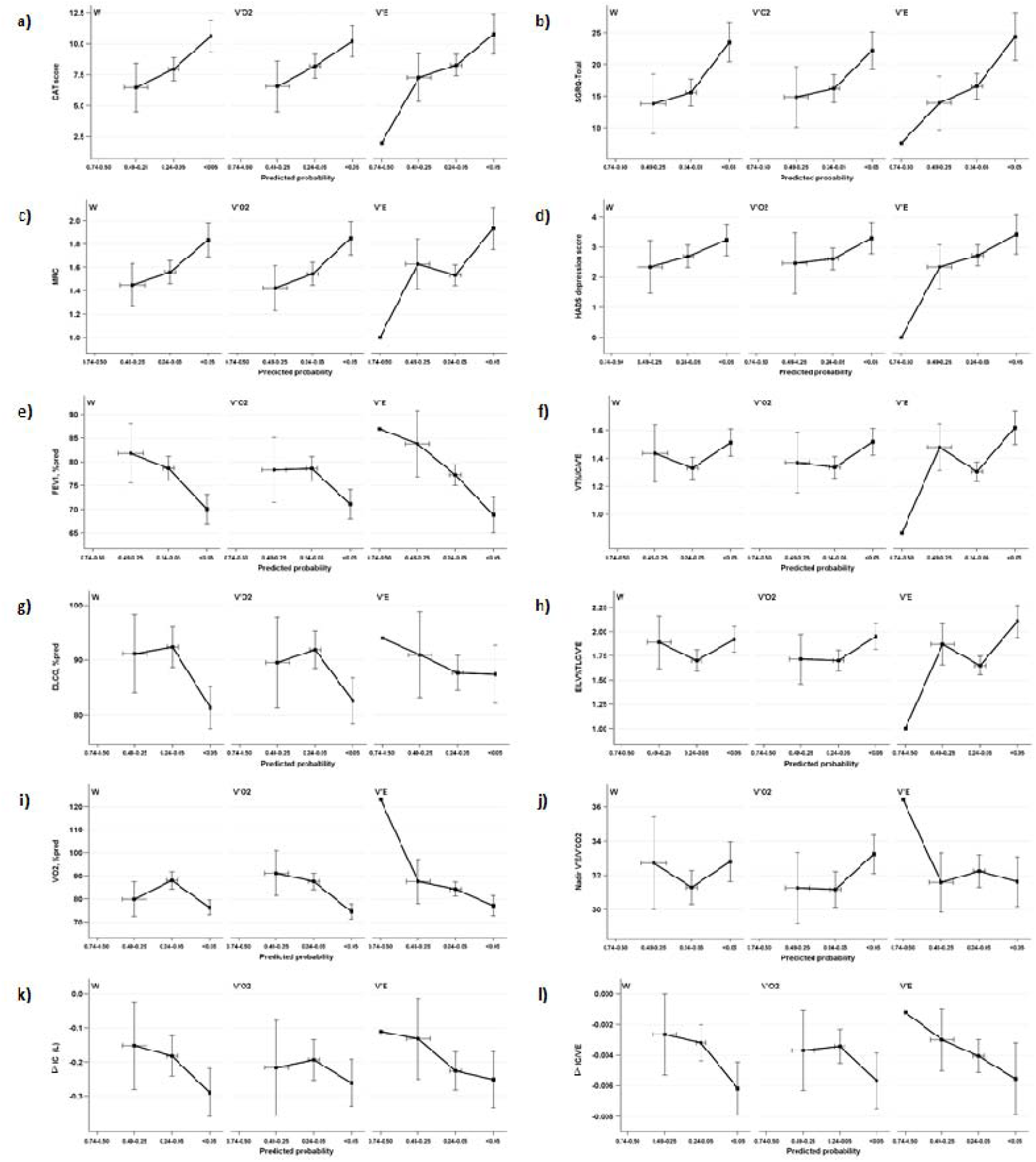
Outcomes by the probability of breathlessness normality, where a lower probability reflects more abnormal breathlessness. The probability was calculated using normative reference equations of the breathlessness intensity response during cycle cardiopulmonary exercise testing (CPET), in relation to power output (W), oxygen uptake (V’O_2_), and minute ventilation (V’_E_) at peak exercise. People with a lower probability of breathlessness normality (more abnormal breathlessness) generally had worse outcomes including lung function, self-reported symptom burden and physical activity, health-related quality of life, exercise capacity, and developing more physiological abnormalities during the CPET.

### Presence of abnormal breathlessness

The prevalence of abnormal breathlessness (Borg CR10 breathlessness intensity rating > ULN) at peak exercise was 22.7% in relation to W, 21.5% in relation to V’O_2_, and 15.2% in relation to V’_E_ (**Table 2**). Overlap between the categorizations of abnormal breathlessness using the different equations is shown in **Figure S2**. Almost all participants with abnormal breathlessness in relation to V’_E_ were also abnormal in relation to V’O_2_, with very few (n=4) participants abnormal relative to V’_E_ only.

**Table 2.**
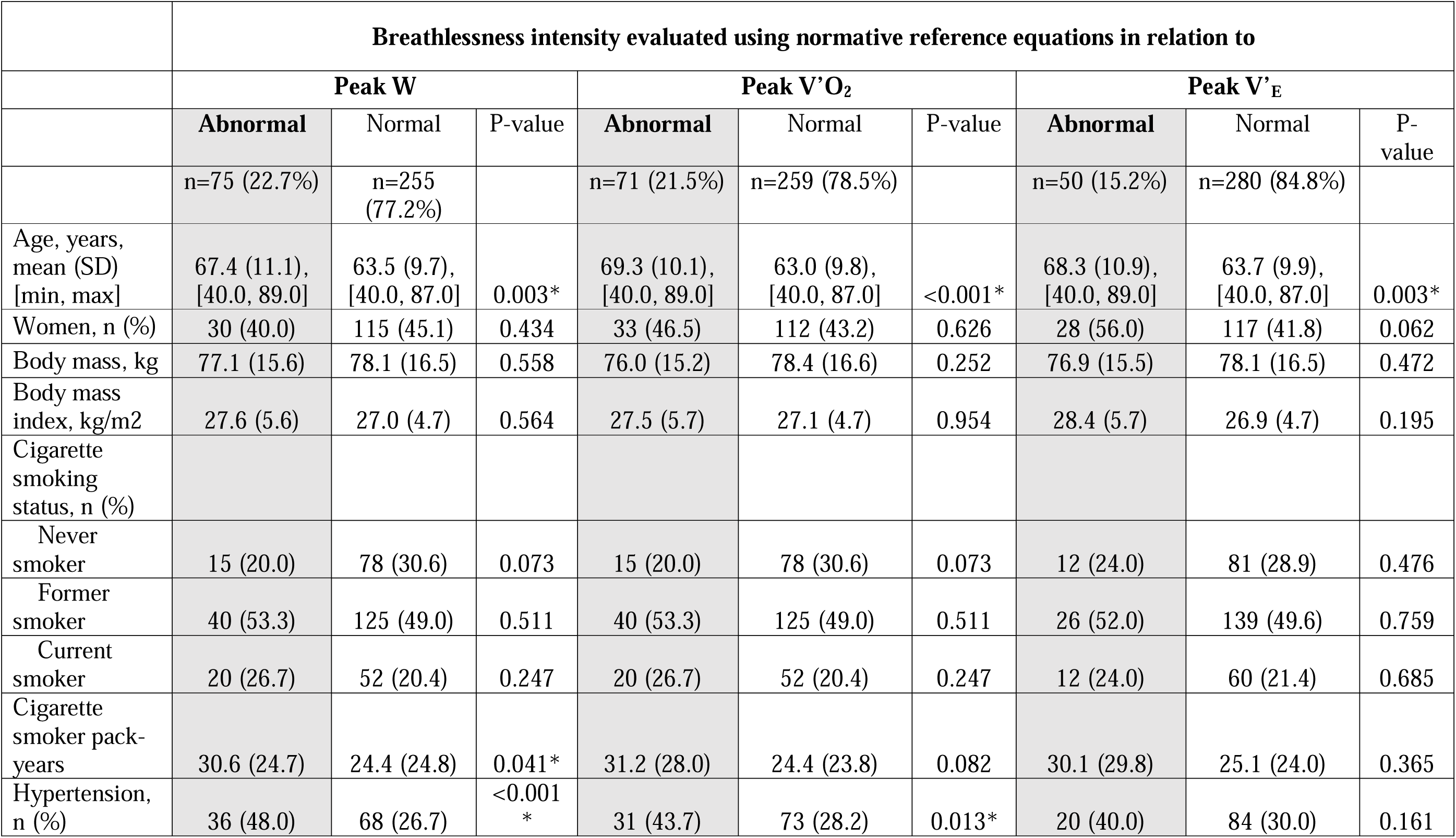
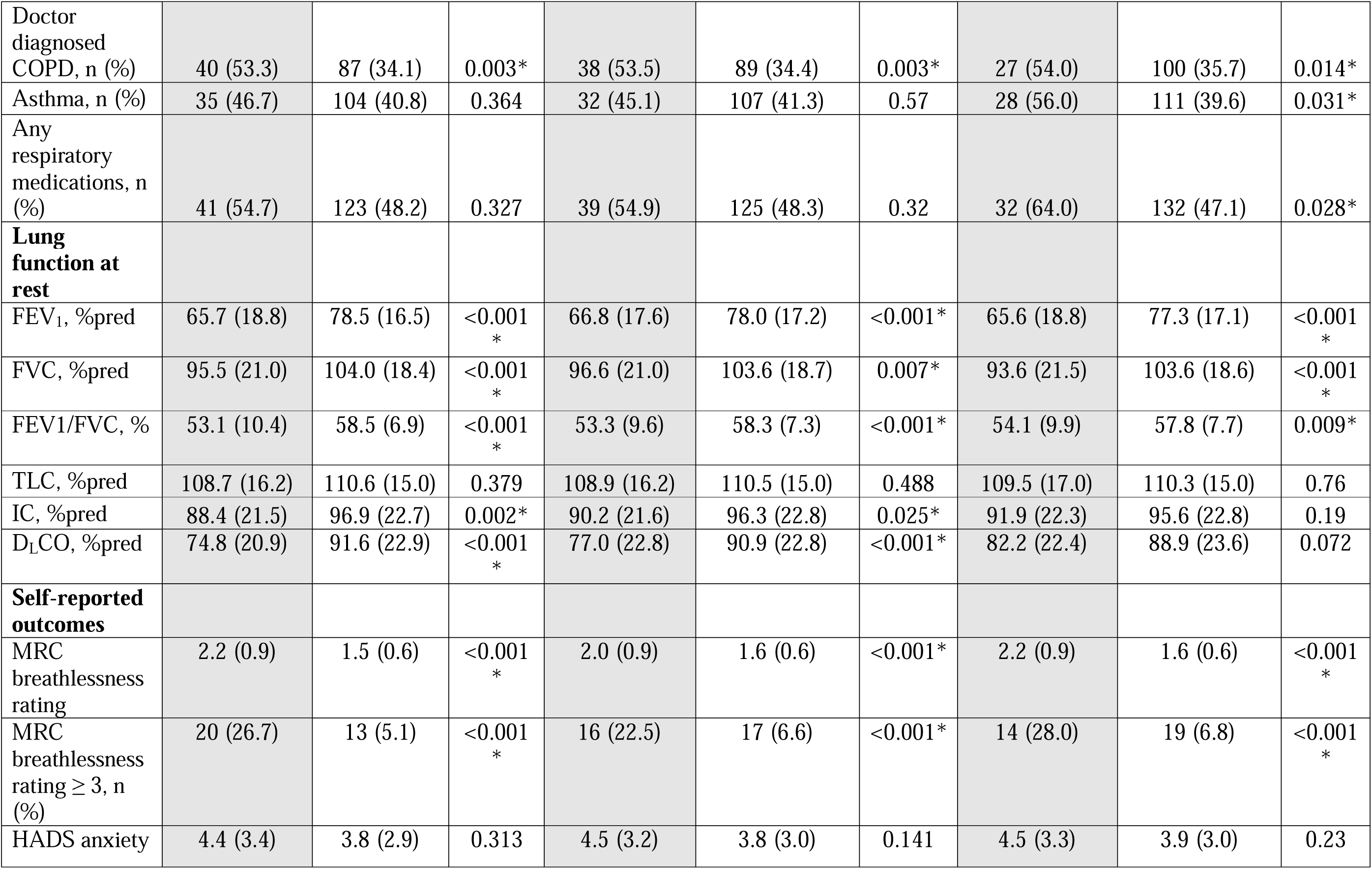
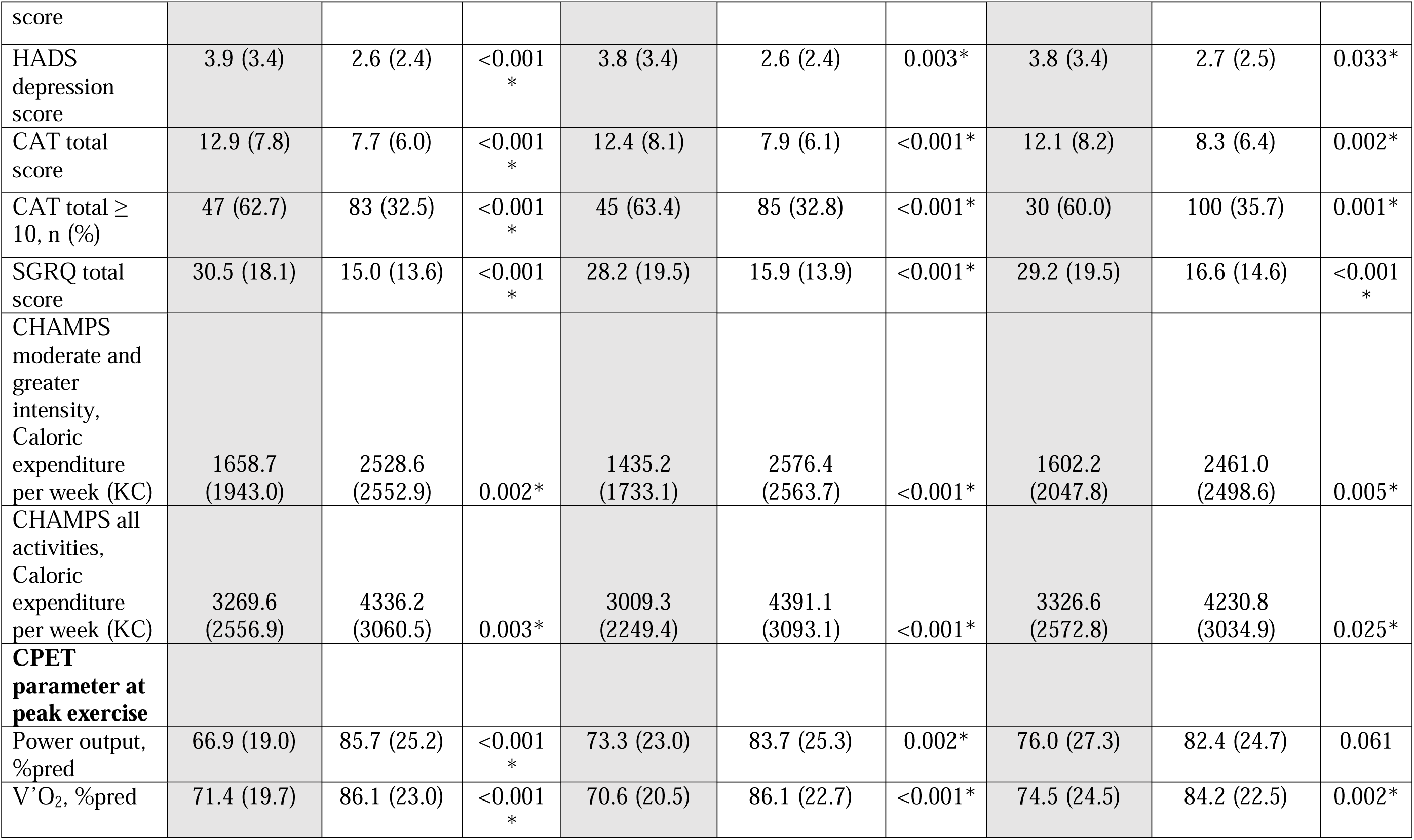
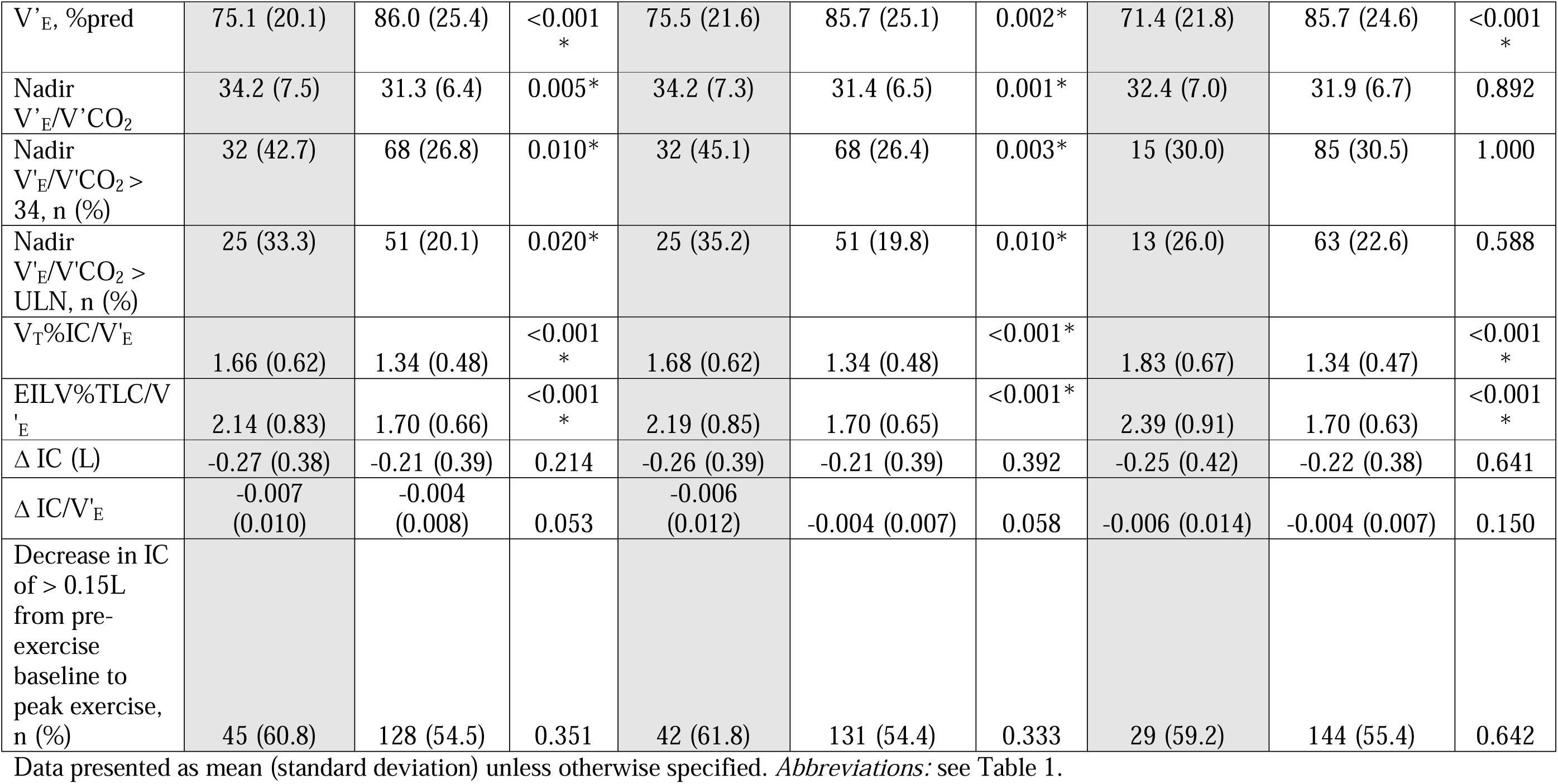
Outcomes in people with abnormal *vs.* normal breathlessness at the symptom-limited peak of incremental cardiopulmonary cycle exercise testing.

#### Discriminative validity

For all three normative reference equations, people with abnormal *vs.* normal exertional breathlessness presented with: more severe chronic airflow limitation (lower FEV_1_/FVC); worse lung function (lower FEV_1_, FVC and D_L_CO %predicted); higher respiratory symptom burden (higher MRC, CAT and SGRQ scores); lower self-reported physical activity; greater exercise intolerance (lower peak W and V’O_2_); and more severe critical inspiratory constraints at peak exercise (higher V_T_%IC/V’_E_ and EILV%TLC/V’_E_) (**Table 2**). In addition, for reference equations using W and V’O_2_ (but not V’_E_), people with abnormal *vs.* normal exertional breathlessness presented with more prevalent and severe exercise ventilatory inefficiency (higher nadir V’_E_/V’CO_2_; and greater proportion of people with nadir V’_E_/V’CO_2_ >34 or >ULN) **(Table 2)**.

#### Concurrent validity

With few exceptions, the correlations between abnormal exertional breathlessness and the outcomes were similar when using the different normative reference equations (**Table S1**). In contrast to abnormal exertional breathlessness in relation to peak W or V’O_2_, having abnormal exertional breathlessness in relation to peak V’_E_ was not correlated with D_L_CO (% predicted), peak W (% predicted) or nadir V’_E_/V’CO_2_ (**Table 2, Table S1**).

## DISCUSSION

This is the first validation of the CPET normative reference equations ^9^ for exertional breathlessness intensity in people with chronic airflow limitation. The main findings are that exertional breathlessness measured as i) the probability of breathlessness normality at peak exercise, and ii) presence of abnormal breathlessness at peak exercise showed discriminative validity by severity of airflow limitation and exercise intolerance, and concurrent validity with other relevant participant-reported and physiological outcomes. In our sample of relatively asymptomatic older adults with mostly mild-to-moderate chronic airflow limitation, people with abnormal breathlessness on CPET had worse lung function, lower exercise capacity, greater respiratory symptom burden, lower self-reported physical activity, and worse HrQoL.

Due to the population-based design, CanCOLD includes a high proportion of relatively healthy participants, with only mild airflow limitation, undiagnosed respiratory disease, no respiratory medication, and no or low burden of self-reported symptoms in daily life ^9,15^. This is likely to explain the relatively low prevalence (<25%) of abnormal breathlessness in this sample. As detection of symptoms is particularly challenging in healthier, asymptomatic populations (in contrast to in people with established, more severe disease), this makes CanCOLD particularly informative for evaluating the validity of methods for breathlessness assessment.

Supporting the validity of the CPET normative reference equations, people with abnormal exertional breathlessness (compared to those with breathlessness within normal ranges) had greater underlying exercise physiological abnormalities known to contribute to exertional breathlessness ^8^. Specifically, people with abnormal *vs.* normal exertional breathlessness in relation to both peak W and V’O_2_ presented with greater exercise ventilatory inefficiency (higher nadir V’_E_/V’CO_2_) and greater critical inspiratory constraints (higher V_T_%IC/V’_E_ and EILV%TLC/V’_E_) at peak exercise, whereas people with abnormal *vs.* normal exertional breathlessness in relation to peak V’_E_ did not have greater exercise ventilatory inefficiency but did present with greater critical inspiratory constraints at peak exercise.

Taken together, the results support that the CPET normative reference equations ^9^ are valid for categorizing the presence and level of abnormal exertional breathlessness in people with chronic airflow limitation. These findings have several important implications. First, they reinforce the use of CPET as the gold standard method to evaluate exertional breathlessness in terms of underlying mechanisms, treatment effects ^8,33-35^, and also the presence and level of abnormal exertional breathlessness. Second, the CPET normative reference equations provide the first valid benchmark for defining abnormal exertional breathlessness. Abnormal breathlessness during CPET can be used to evaluate and compare different ways of assessing breathlessness and/or respiratory symptom burden such as task-based questionnaires like MRC and CAT; to select participants for clinical (therapeutic) trials based on the presence and/or level of abnormal exertional breathlessness. The current practice to determine breathlessness severity and trial eligibility using questionnaires such as the modified MRC is limited by misinterpretation ^36,37^, misclassification ^38^, and failure to evaluate breathlessness at a standardized level of exertion or ventilation, which may lead to breathlessness being underreported or hidden in people who have restricted their physical activity to avoid undue breathlessness ^5,39^. These problems are likely to be overcome by categorizing the level of exertional breathlessness using CPET ^39,40^. Third, we have validated all three normative reference equations to use when evaluating an individual’s exertional breathlessness response – in relation to W, V’O_2_ and V’_E_. In line with our current understanding of the pathophysiological mechanisms of exertional breathlessness in people with chronic airflow limitation ^8^, breathlessness intensity ratings that were abnormal in relation to peak W and/or V’O_2_ were accompanied by evidence of (i) more prevalent and severe exercise ventilatory inefficiency and (ii) more severe critical inspiratory constraints, whereas breathlessness intensity ratings that were abnormal in relation to peak V’_E_ were associated with more severe critical inspiratory constraints only. An intriguing next step is to see if an individual’s profile of exertional breathlessness abnormality (by V’_E_ *and/or* V’O_2_) might help to identify the underlying physiological mechanism(s) contributing to abnormal exertional breathlessness in clinical practice.

### Which of the different reference equations should be used to evaluate the (ab)normality of exertional breathlessness?

Based on the present findings, we suggest that evaluation of exertional breathlessness in relation to both peak V’O_2_ and peak V’_E_ should be the first line option in most situations. This is based on the high degree of overlap between categorization exertional breathlessness by peak W and peak V’O_2_ (both measures of exercise intensity or level of exertion), and that the equation by peak V’O_2_ identified all participants who were also abnormal by peak V’_E_. Defining abnormal exertional breathlessness using reference equations for both peak W and peak V’O_2_ could, however, identify some additional people as having abnormal exertional breathlessness. The cross-talk between the different reference equations to categorize the (ab)normality of exertional breathlessness needs to be further explored.

Strengths of this study is the multicenter, population-based design, which reduces the risk of selection bias; the wide range of relevant outcome measures, including self-reported respiratory symptom burden and HrQoL using validated questionnaires (MRC, CAT, SGRQ), lung function (spirometry, plethysmography, D_L_CO), and detailed physiological outcomes assessed during symptom-limited incremental cycle CPET – a breadth of factors unique to the CanCOLD dataset ^15^ and that are optimal for demonstrating both the discriminative and concurrent validity of the CPET normative reference equations for exertional breathlessness ^9^.

Next research steps include validating the normative reference equations in people with more severe chronic airflow limitation, and in other chronic health conditions such as interstitial lung disease and heart disease. Further research possibilities include: evaluating the prevalence of abnormal exertional breathlessness across populations and patient groups; defining the method’s responsiveness and minimal clinically important difference; and to evaluate the prognostic implication of abnormal exertional breathlessness on clinical outcomes and risk of premature death.

In conclusion, evaluation of the presence and level of abnormal breathlessness using CPET normative reference equations (in relation to peak W, V’O_2_, or V’_E_) showed discriminative and concurrent validity in relation to other relevant clinical and patient-reported outcomes, including lung function, respiratory symptom burden, self-reported physical activity, HrQoL, exercise capacity, and underlying exercise physiologic abnormalities among people with chronic airflow limitation. Through use of these normative reference equations, researchers and clinicians can, for the first time, evaluate the presence and quantify the level of abnormally high exertional breathlessness, in patient groups or in the individual.

## Data Availability

The data used in the present study cannot be freely shared due to Canadian legal and ethical restrictions, but data may be shared upon reasonable request to the authors and after separate ethical approval.

## Acknowledgments

The authors thank the people who participated in the study and the many members of the CanCOLD collaborative research group: **Executive Committee:** Jean Bourbeau (McGill University, Montreal, QC, Canada); Wan C Tan, J Mark FitzGerald, Don D Sin (University of British Columbia, Vancouver, BC, Canada); Darcy D Marciniuk (University of Saskatoon, Saskatoon, SK, Canada); Denis E O’Donnell (Queen’s University, Kingston, ON, Canada); Paul Hernandez (Dalhousie University, Halifax, NS, Canada); Kenneth R Chapman (University of Toronto, Toronto, ON, Canada); Brandie Walker (University of Calgary, Calgary, AB, Canada); Shawn Aaron (University of Ottawa, Ottawa, ON, Canada); François Maltais (University of Laval, Quebec City, QC, Canada). **International Advisory Board:** Jonathon Samet (the Keck School of Medicine of USC, California, USA); Milo Puhan (John Hopkins School of Public Health, Baltimore, USA); Qutayba Hamid (McGill University, Montreal, QC, Canada); James C Hogg (University of British Columbia, Vancouver, BC, Canada). **Operations Center:** Jean Bourbeau (Principal Investigator), Dany Doiron, Palmina Mancino, Pei Zhi Li, Dennis Jensen, Carolyn Baglole (McGill University, Montreal, QC, Canada); Yvan Fortier (Laboratoire telematique, Quebec Respiratory Health Network, Fonds de la recherche en santé du Québec (FRQS)); Wan C Tan (co-Principal Investigator), Don Sin, Julia Yang, Jeremy Road, Joe Comeau, Adrian Png, Kyle Johnson, Harvey Coxson, Jonathon Leipsic, Cameron Hague (University of British Columbia, Vancouver, BC, Canada), Miranda Kirby (Ryerson University, Toronto, ON, Canada) **Economic Core:** Mohsen Sadatsafavi (University of British Columbia, Vancouver, BC, Canada). **Public Health Core:** Teresa To, Andrea Gershon (University of Toronto, Toronto, ON, Canada). **Data management and Quality Control:** Wan C Tan, Harvey Coxson (University of British Columbia, Vancouver, BC, Canada); Jean Bourbeau, Pei-Zhi Li, Zhi Song, Andrea Benedetti, Dennis Jensen (McGill University, Montreal, QC, Canada); Yvan Fortier (Laboratoire telematique, Quebec Respiratory Health Network, FRQS); Miranda Kirby (Ryerson University, Toronto, ON, Canada). **Field Centers:** Wan C Tan (Principal Investigator), Christine Lo, Sarah Cheng, Elena Un, Cynthia Fung, Wen Tiang Wang, Liyun Zheng, Faize Faroon, Olga Radivojevic, Sally Chung, Carl Zou (University of British Columbia, Vancouver, BC, Canada); Jean Bourbeau (Principal Investigator), Palmina Mancino, Jacinthe Baril, Laura Labonte (McGill University, Montreal, QC, Canada); Kenneth Chapman (Principal Investigator), Patricia McClean, Nadeen Audisho (University of Toronto, Toronto, ON, Canada); Brandie Walker (Principal Investigator), Curtis Dumonceaux, Lisette Machado (University of Calgary, Calgary, AB, Canada); Paul Hernandez (Principal Investigator), Scott Fulton, Kristen Osterling, Denise Wigerius (University of Halifax, Halifax, NS, Canada); Shawn Aaron (Principal Investigator), Kathy Vandemheen, Gay Pratt, Amanda Bergeron (University of Ottawa, Ottawa, ON, Canada); Denis O’Donnell (Principal Investigator), Matthew McNeil, Kate Whelan (Queen’s University, Kingston, ON, Canada); François Maltais (Principal Investigator), Cynthia Brouillard (University of Laval, Quebec City, QC, Canada); Darcy Marciniuk (Principal Investigator), Ron Clemens, Janet Baran, Candice Leuschen (University of Saskatoon, Saskatoon, SK, Canada).

## Author contributions statement

Study conception and design (ME, HL, DJ); data collection (JB, WCT, DJ); statistical analysis (PZL); first draft (ME); interpretation, revision the manuscript for intellectual contents, and approval of the final version to submit (all authors).

