## Supplemental material for "Abnormal breathlessness during cardiopulmonary exercise testing - validation in people with chronic airflow limitation"

**Table S1.** Correlations between having abnormal breathlessness and outcomes

|  | **Normative reference equation for breathlessness intensity** | | | | | |
| --- | --- | --- | --- | --- | --- | --- |
|  | **Peak W** |  | **Peak V’O_2_** | | **Peak V’_E_** | |
|  | **Point biserial Correlation** | **P value** | **Point biserial Correlation** | **P value** | **Point biserial Correlation** | **P value** |
| **Baseline lung function** |  |  |  |  |  |  |
| FEV_1_, %pred | -0.30 | <0.001 | -0.26 | <0.001 | -0.24 | <0.001 |
| FVC, %pred | -0.18 | <0.001 | -0.15 | 0.007 | -0.19 | <0.001 |
| FEV_1_/FVC, % | -0.28 | <0.001 | -0.26 | <0.001 | -0.17 | 0.002 |
| D_L_CO, %pred | -0.30 | <0.001 | -0.24 | <0.001 | -0.1 | 0.069 |
| **Self-reported outcomes** |  |  |  |  |  |  |
| MRC breathlessness rating | 0.36 | <0.001 | 0.26 | <0.001 | 0.32 | <0.001 |
| HADS anxiety score | 0.08 | 0.155 | 0.08 | 0.125 | 0.07 | 0.185 |
| HADS depression score | 0.21 | <0.001 | 0.19 | <0.001 | 0.14 | 0.011 |
| CAT total score | 0.32 | <0.001 | 0.27 | <0.001 | 0.2 | <0.001 |
| SGRQ-Total score | 0.40 | <0.001 | 0.32 | <0.001 | 0.28 | <0.001 |
| CHAMPS moderate and greater intensity, Caloric expenditure per week (KC) | -0.15 | 0.008 | -0.19 | <0.001 | -0.12 | 0.023 |
| CHAMPS all activities, Caloric expenditure per week (KC) | -0.15 | 0.007 | -0.19 | <0.001 | -0.11 | 0.05 |
| **Symptom-limited peak CPET parameters** |  |  |  |  |  |  |
| Peak power output, %pred | -0.31 | <0.001 | -0.17 | 0.002 | -0.09 | 0.099 |
| Peak V’O_2_, %pred | -0.27 | <0.001 | -0.28 | <0.001 | -0.15 | 0.006 |
| Peak V’_E_, %pred | -0.19 | <0.001 | -0.17 | 0.002 | -0.21 | <0.001 |
| Nadir V’_E_/V’CO_2_ | 0.18 | <0.001 | 0.17 | 0.002 | 0.02 | 0.682 |
| Peak V_T_%IC/V'_E_ | 0.26 | <0.001 | 0.27 | <0.001 | 0.34 | <0.001 |
| Peak EILV%TLC/V'_E_ | 0.25 | <0.001 | 0.28 | <0.001 | 0.35 | <0.001 |

Abnormal breathlessness defined as a breathlessness (Borg CR10) intensity rating above the predicted upper limit of normal (ULN) at peak exercise. The ULN was calculated using normative reference equations in relation to peak power output (W), oxygen uptake (V’O_2_), and minute ventilation (V’_E_). *Abbreviations:* see Table 1.

**Figure S1.** Distribution of the predicted probability for breathlessness normality at peak exercise during incremental cycle cardiopulmonary exercise testing (CPET), in a population-based sample (N=330) of people with chronic airflow limitation. A lower predicted probability reflects more abnormal (severe) breathlessness.

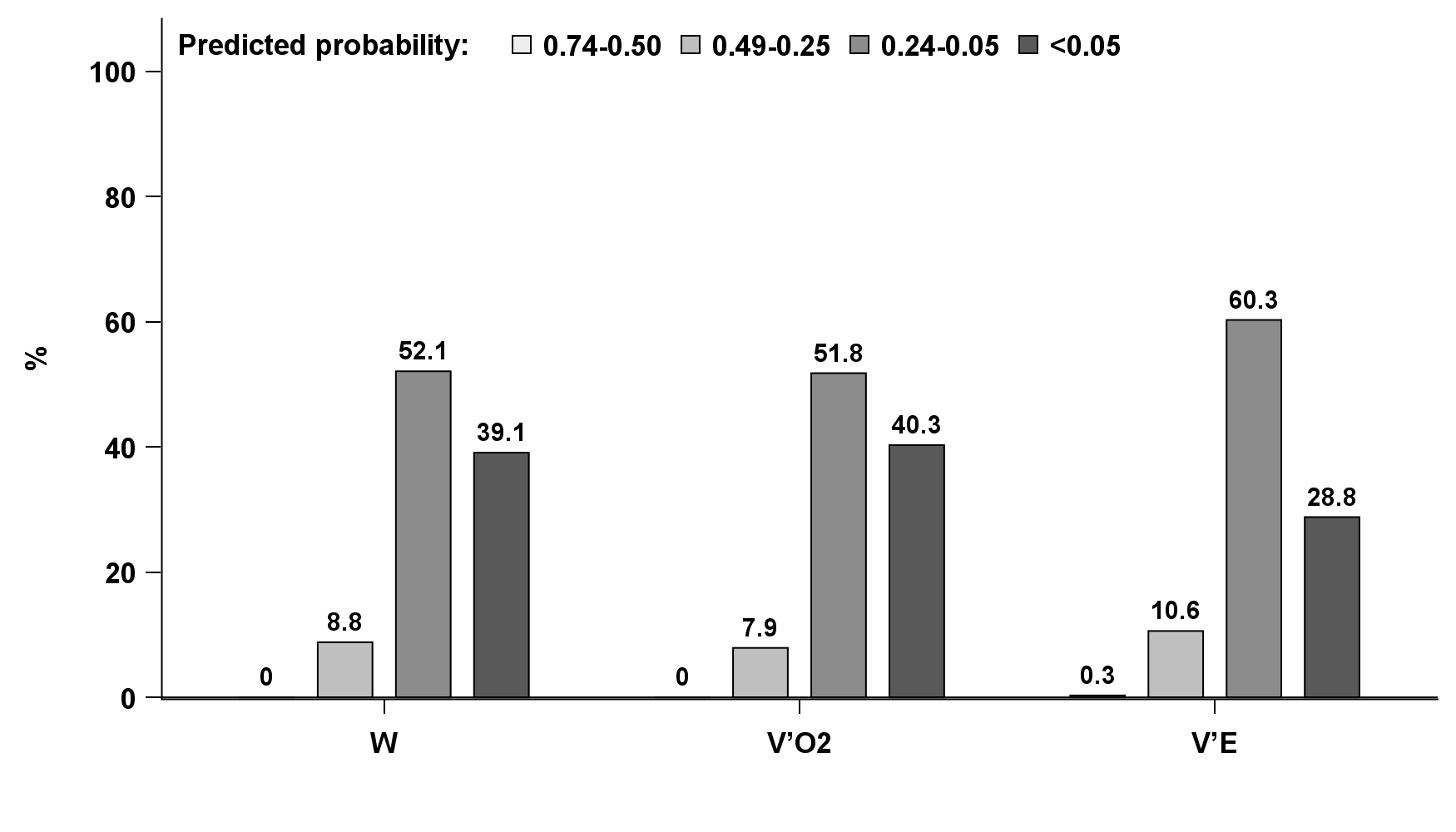

**Figure S2.** The overlap between categorization of presence of abnormal breathlessness using normative reference equations, in relation to power output (W), rate of oxygen uptake (V’O_2_), and minute ventilation (V’_E_) in 330 people with chronic airflow limitation.

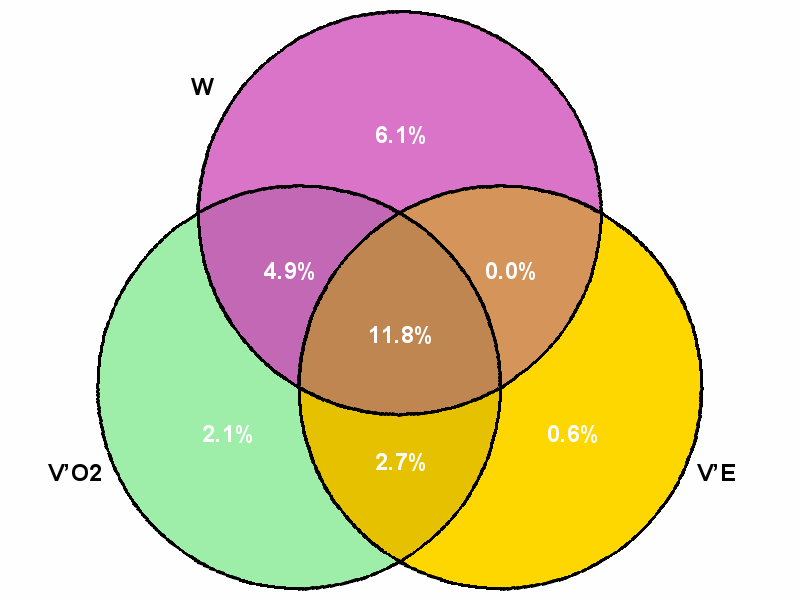
